# A scalable saliva-based, extraction-free rt-lamp protocol for sars-cov-2 diagnosis

**DOI:** 10.1101/2020.10.27.20220541

**Authors:** Paula Asprino, Fabiana Bettoni, Anamaria Camargo, Diego Coelho, Guilherme Coppini, Igor Correa, Erika Freitas, Lilian Inoue, João Paulo Kitajima, Mayra Kuroki, Cibele Masotti, Tatiana Marques, Alice Reis, Luiz Fernando Reis, Bibiana Santos, Ernande dos Santos, David Schlesinger, Cecília Sena, Talita Spadaccini, Lucas Taniguti

## Abstract

Scalable, cost-effective screening methods are an essential tool to control SARS-CoV-2 spread. We have developed a straight saliva-based, RNA extraction-free, RT-LAMP test that is comparable to current nasopharyngeal swab RT-PCR tests in both sensitivity and specificity. Using a 2-step readout of fluorescence and melting-point curve analysis, the test is scalable to more than 30,000 tests per day with average turnaround time of less than 3 hours. The test was validated using samples from 244 symptomatic patients, and showed sensitivity of 78.9% (vs. 85.5% for nasopharyngeal swabs RT-PCR) and specificity of 100% (vs. 100% for nasopharyngeal swabs RT-PCR). Our method is therefore accurate, robust, time and cost effective and therefore can be used for screening of SARS-CoV-2.

## II. INTRODUCTION

The corona virus disease 2019 (COVID-19) caused by the SARS-CoV-2 (Severe acute respiratory syndrome coronavirus 2) has become a major public health emergency worldwide. Accurate diagnosis of COVID-19 is crucial to control disease transmission and manage infected patients.

Reverse transcription followed by real-time Polymerase chain reaction (RT-PCR) assays using RNA extracted from nasopharyngeal swabs are the gold standard for SARS-CoV-2 molecular diagnostic and are currently the screening strategy used worldwide. These methods are cost and labor-intensive, including several steps that require sample handling, and have been restrained by lack of real-time PCR-specific instruments and reagents.

Sample collection through nasopharyngeal swabs itself is also a major barrier to high-scale, low-cost testing due to limited availability of swabs and necessity of a trained professional, themselves at risk for infection. In this context, saliva has emerged as a viable non-invasive alternative for viral detection, showing comparable viral loads for SARS-CoV-2 in infected patients. (Anne L. Wyllie et al. 2020; Anne Louise Wyllie et al. 2020). Saliva auto-collection is an enticing proposition that simplifies sample collection logistics and reduces infection risk of healthcare workers.

RNA extraction is another important bottleneck for COVID-19 diagnosis. Supply of reagents and automation hardware are limited and incompatible with the worldwide demand. Single-step RNA extraction solutions increase sensitivity but add complexity and cost to the process.

In 2000, Notomi et al. developed a DNA amplification method based on an isothermal reaction: Loop-mediated isothermal amplification (LAMP). (Notomi et al. 2000) LAMP is faster than PCR, utilizes a different DNA polymerase, and can be read out through multiple methods, ranging from simple colorimetric (Zhang et al. 2020), to direct fluorescence (Lu et al. 2020), Crispr-based assays (Joung et al. 2020), and sequencing (James et al., n.d.). LAMP methodology has been used for diagnosis of influenza virus (Poon et al. 2005), Ebola virus (Kurosaki, Magassouba, and Oloniniyi 2016), and a variety of other pathogens (Bartolone et al. 2018).

Several groups have published LAMP protocols for SARS-CoV-2 diagnosis, including FDA-approved protocols. However, the majority of these protocols are based on direct nasopharyngeal swab samples followed by RNA extraction which are still associated with some limitations, as has been discussed previously.

To contribute towards solutions for Covid-19 diagnosis, we combined and optimized previously published protocols to establish a simplified, saliva-based, RNA extraction-free RT-LAMP test that centralized labs can execute at large-scale population levels.(Lalli, Langmade, et al. 2020) By focusing on simplicity as the primary goal, we have added to this body of scientific literature by creating a test that is cheap, fast, scalable, and accurate.

## III. MATERIALS AND METHODS

### A. Subjects

IRB approval (HSL 2020-43) was obtained prior to clinical studies. Subjects included in the study presented to the Hospital Sírio-Libanês in São Paulo, Brazil with 1-7 days of symptoms suggestive of Covid-19. All subjects were diagnosed by routine nasopharyngeal swab RT-PCR collected at the same time as the saliva samples.

### B. Sample Collection

Saliva samples were collected by patients themselves in sterile 50 ml conical tubes with no additives. Approximately 1 ml of saliva was collected and the pre-collection instruction was to refrain from eating, drinking or smoking for 60 minutes. Samples were then stored at room temperature for a period ranging from 1 to 3 days.

### C. RT-LAMP

The RNA-extraction free protocol for RT-LAMP was adapted from (Lalli, Chen, et al. 2020; Lamb et al. 2020). Saliva aliquots (50 ul) were pre-heated at 98oC for 10 minutes to release RNA from cells and 8ul of the supernatant were transferred to 42ul of RT-LAMP reaction mix. The reaction was incubated for 5 minutes at room temperature, for Uracil-DNA-Glycosylase (UDG) to eliminate eventual carryover contaminants. Using a conventional thermocycler, RT-LAMP reaction (table 1) was heated to 63oC for 30 minutes and then reaction was stopped by heating to 80oC for an additional 5 minutes. Primer set (table 3) consists of six primers designed by Joung and colleagues. (Joung et al. 2020) to target the Nucleocapsid gene, which has higher copy numbers than other segments of the SARS-CoV-2 genome. Synthetic SARS-CoV-2 RNA from Twist Biosciences was used as positive control and nuclease-free water and saliva from **healthy**subjects as negative controls.

**Table 1:**
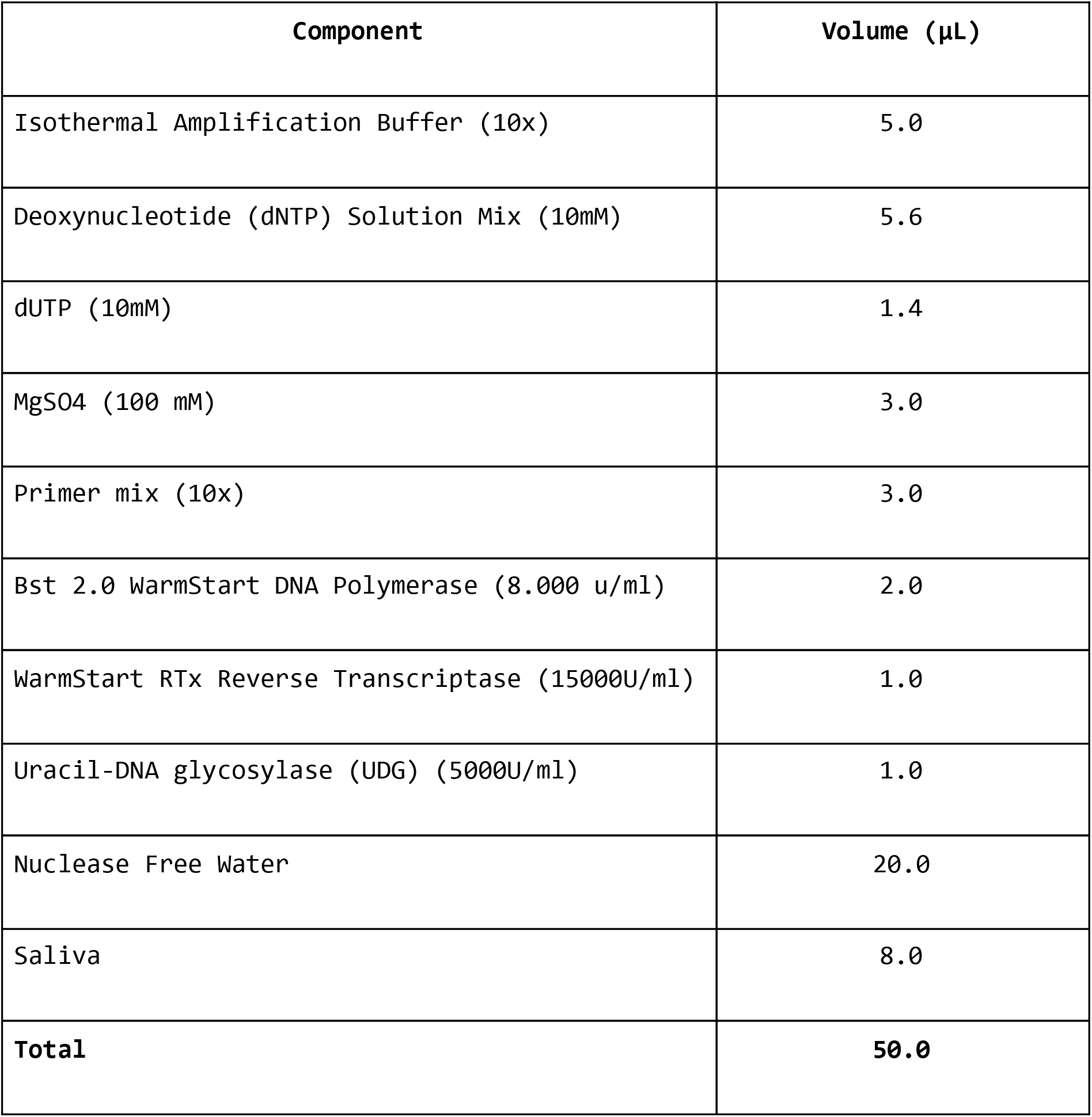
RT-LAMP Mix

| Component | Volume (µL) |
| --- | --- |
| Isothermal Amplification Buffer (10x) | 5.0 |
| Deoxynucleotide (dNTP) Solution Mix (10mM) | 5.6 |
| dUTP (10mM) | 1.4 |
| MgSO<sub>4</sub> (100 mM) | 3.0 |
| Primer mix (10x) | 3.0 |
| Bst 2.0 WarmStart DNA Polymerase (8.000 u/ml) | 2.0 |
| WarmStart RTx Reverse Transcriptase (15000U/ml) | 1.0 |
| Uracil-DNA glycosylase (UDG) (5000U/ml) | 1.0 |
| Nuclease Free Water | 20.0 |
| Saliva | 8.0 |
| **Total** | **50.0** |

**Table 2:** Primer Mix

| **Volume (μL)** | **Primer** | **Initial**<br>**Concentration (μM)** | **Final**<br>**Concentration**<br>**(μM)** |
| --- | --- | --- | --- |
| 80 | FIP(F1c+F2)_RT-LAMP | 100 | 16 |
| 80 | BIP(B1c+B2)_RT-LAMP | 100 | 16 |
| 10 | F3_RT-LAMP | 100 | 2 |
| 10 | B3_RT-LAMP | 100 | 2 |
| 20 | LoopF_RT-LAMP | 100 | 4 |
| 20 | LoopB_RT-LAMP | 100 | 4 |
| 280 | H2O | - | - |
| 500 | **Total** |  |  |

**Table 3:**
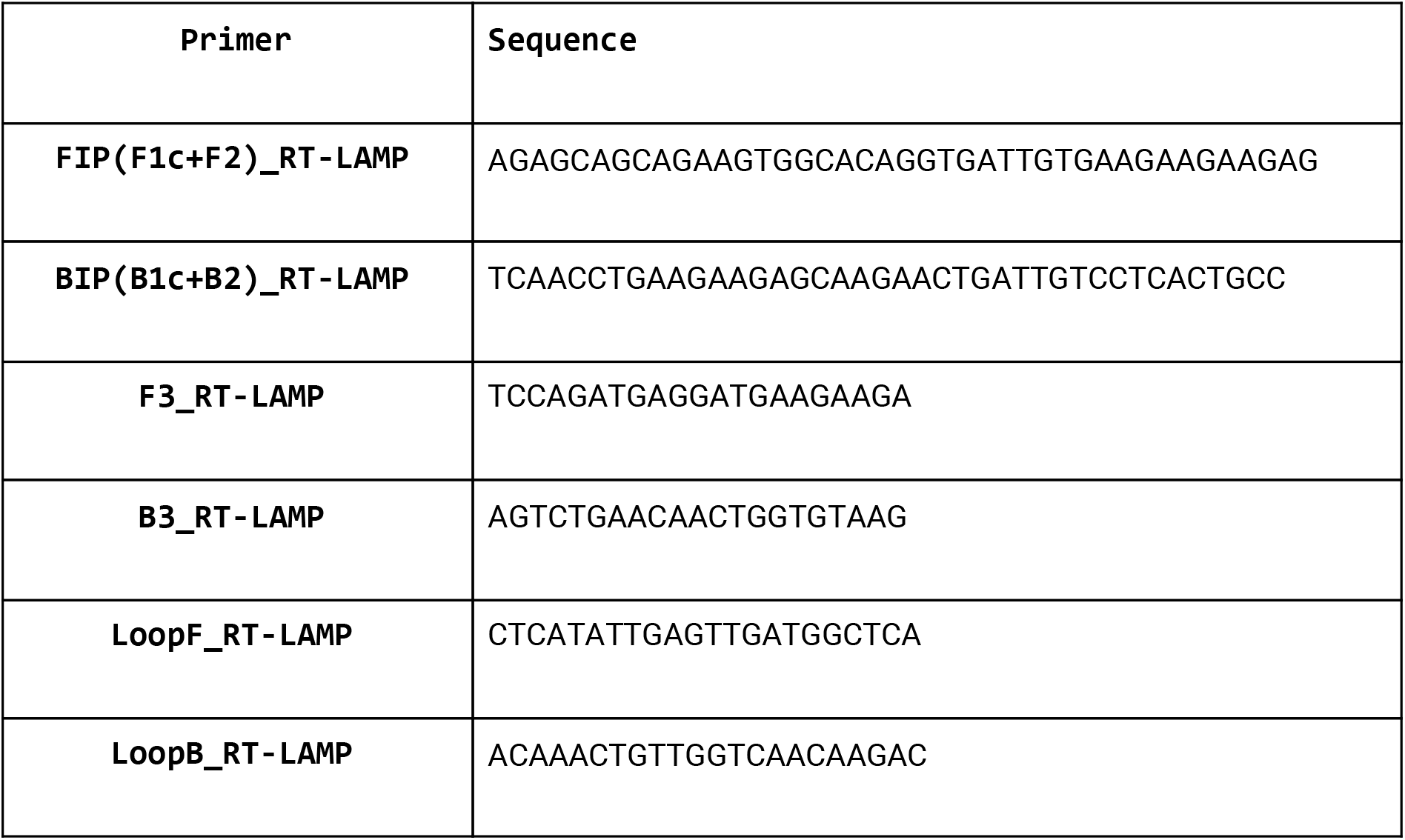
Primer Sequences

### D. RT-LAMP Readout

RT-LAMP products were diluted 40x with EvaGreen fluorescent dye (CellCo, 100x diluted in TAE1x) and read in a fluorescence spectrophotometer. RT-LAMP products from positive samples were also analyzed by Agilent 2100 Bioanalyzer and real time PCR equipment.

Readings in the fluorescence spectrophotometer (Promega Glomax) were considered negative if the diluted reaction had less than 1500 RFU/ul. The fluorescence in each well was measured once, with excitation modules and emission at 475nm and 500–550nm respectively.

Resulting RT-LAMP products were also analyzed using Bioanalyzer, for characteristic banding patterns (Lamb et al. 2020) and confirmed by dissociation curve analysis in a real time PCR equipment (CFX96 Touch Real-Time PCR Thermal Cycler Biorad or 7300-Applied Biosystems), using cycling parameters as follows: 63°C for 1:00 min, 95°C for 15 seconds, melt analysis performed from 63°C to 95°C, in a method broadly similar to (Rolando et al. 2020). The incorporation of dissociation curves allows the confirmation of low viral load positives from negatives samples, as well as increases the specificity for SARS-CoV-2 detection.

## IV. RESULTS

### A. ANALYTICAL SENSITIVITY

RT-LAMP analytical sensitivity was determined by limiting dilution of synthetic SARS-CoV-2 RNA (Twist Bioscience). Ten replicates of six different concentrations ranging from 10,000 copies to 10 copies per reaction were processed through the assay. Limit of detection (LoD) was defined as the lowest concentration at which >90% of the replicates were detected. LoD was determined as 20 copies per reaction (cutoff −d(RFU)/dT = 20). (**Figure 1**). Considering the use of 8 μl of saliva as input, the estimated detection limit is 2.5 copies per μl.

**Figure 1:**
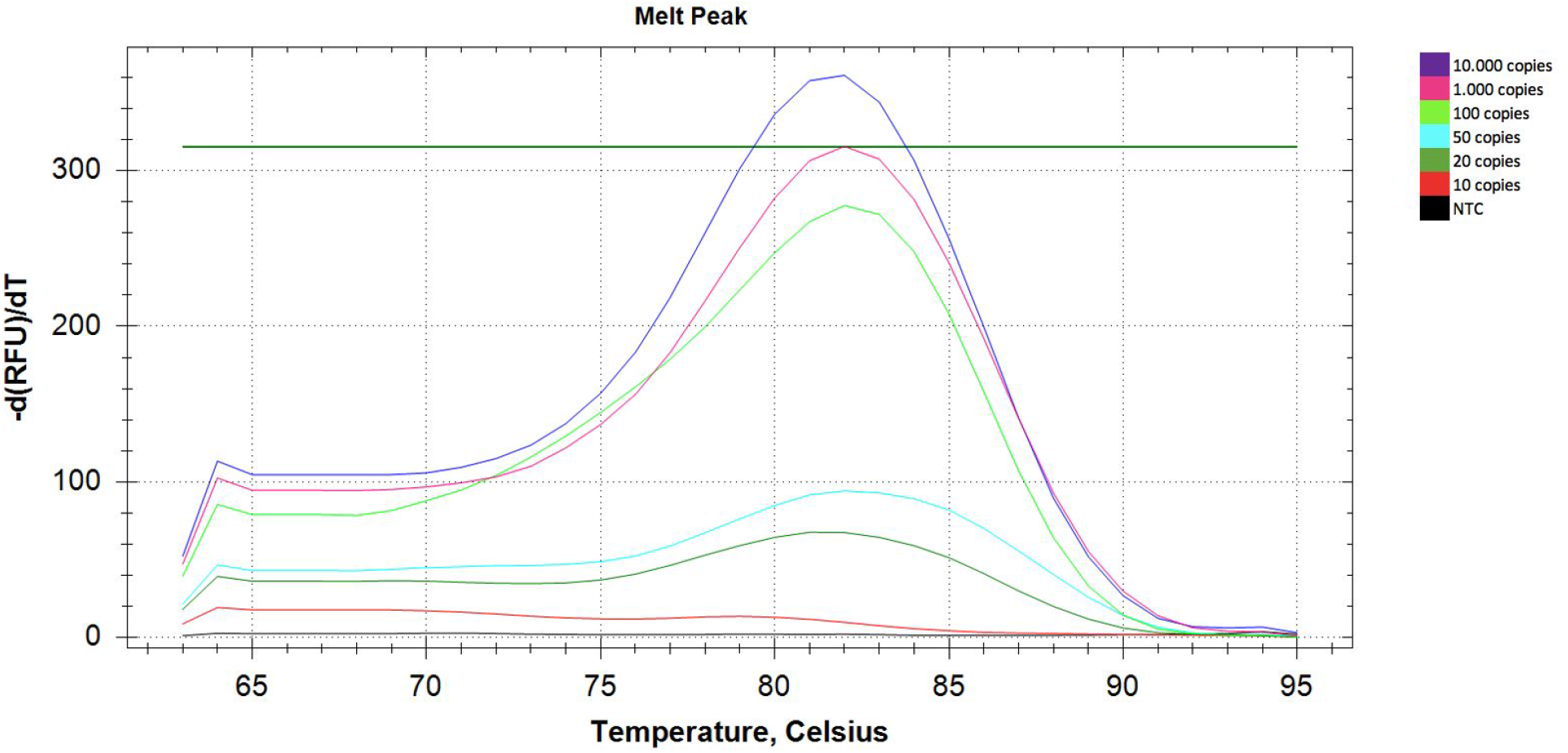
Dissociation curves observed for positive samples in a real-time pcr equipment. These curves permit to increase the sensitivity for low viral load samples and guarantees calling of true positives, conferring higher specificity.

### B. REPLICABILITY

Inter-plate replicability was determined using 122 consecutive plates containing 1 positive control and 1 negative control. (Figure 2). All 122 positive controls scored above the 1,900 RFU stringent cutoff (minimum positive control value = 2,066 RFU). All 122 negative controls scored below the more sensitive cutoff of 1,500 used for subsequent melting point evaluation (maximum negative control value = 817 RFU). In Figure 3, 50,000 consecutive samples, positive and negative subjects show a similar fluorescence readout distribution to corresponding controls.

**Figure 2:**
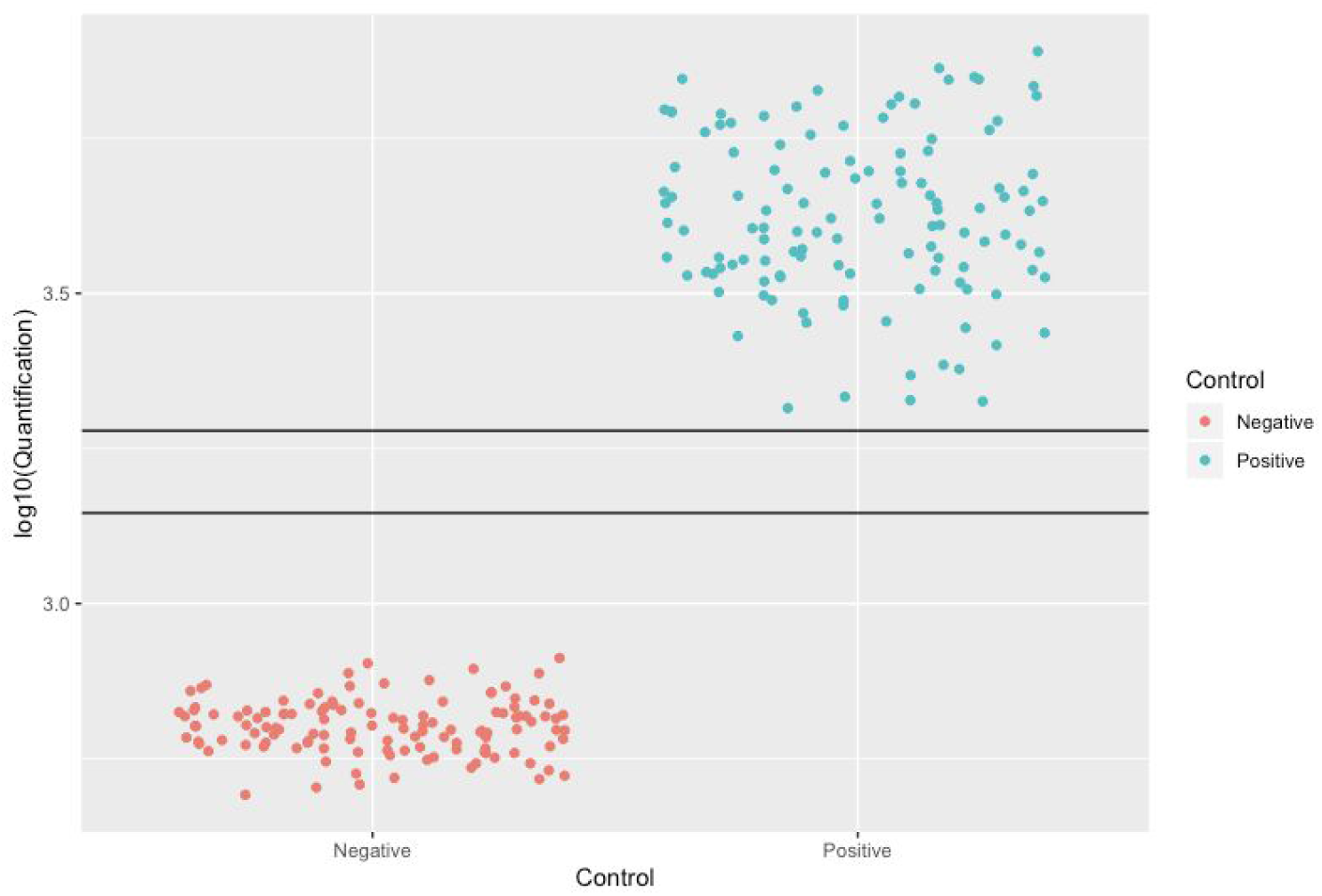
Positive and negative control samples from multiple reactions show fluorescence replicability.

**Figure 3:**
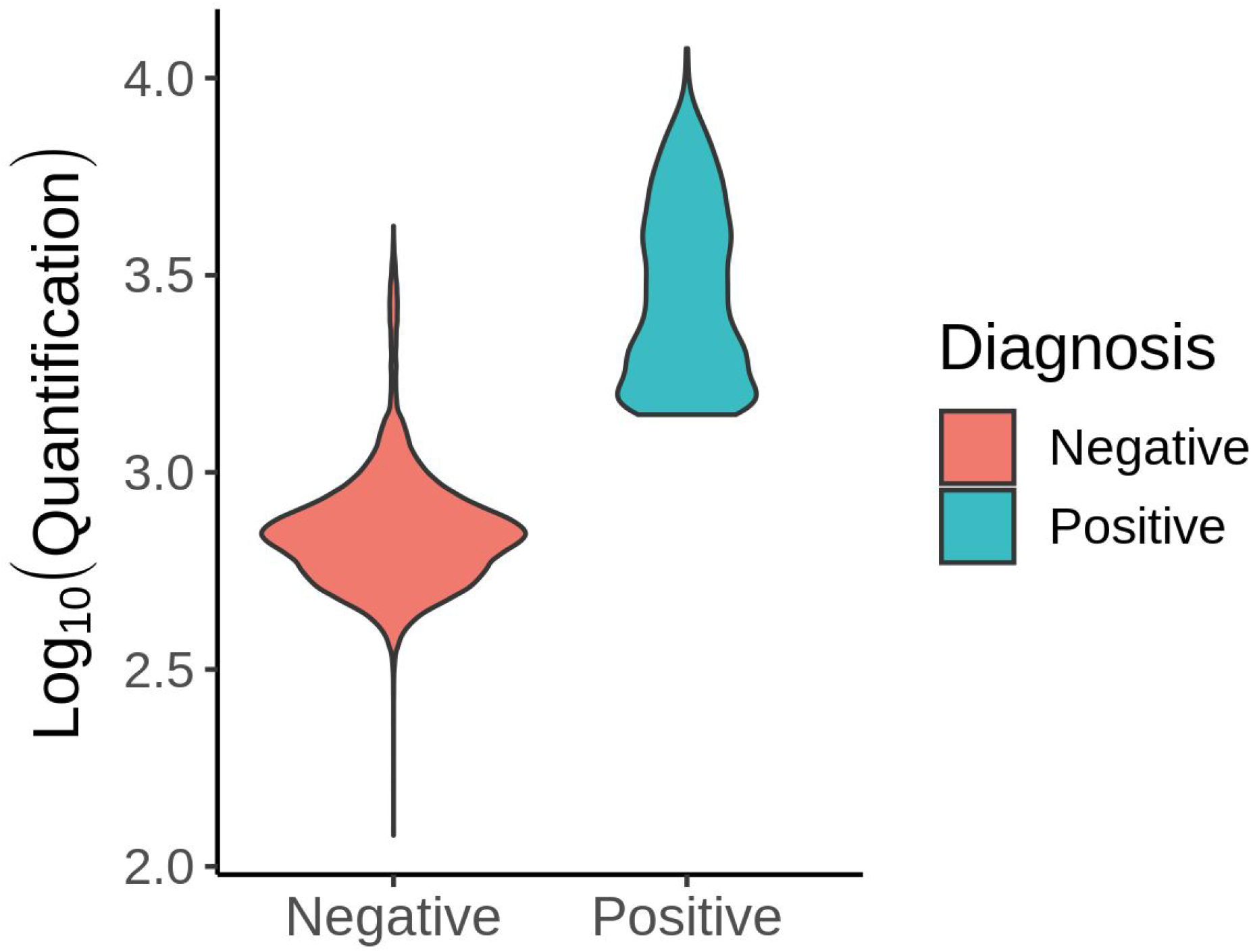
LAMP Results Fluorescence Distribution of 50,000 consecutive saliva samples.

### C. CLINICAL PERFORMANCE

Saliva RT-LAMP results were compared to nasopharyngeal RT-PCR results from an independent lab. Validation was independently carried out by Mendelics and Hospital Sírio-Libanês, and the results were combined. RT-LAMP and RT-PCR results were blinded to the complementary results.

As shown in Table 4, a total of 244 symptomatic patients, ranging from days 1 through 7 of symptoms, were tested. SARS-CoV-2 RNA was detected in 31% (76/244) of patients by at least one method. RT-PCR identified 65 of 76 patients (sensitivity of 85.5%) while RT-LAMP identified 60 of 76 patients (sensitivity of 78.9%). Sixteen positive patients were identified by RT-PCR only, while eleven by RT-LAMP only. Specificity was 100% for both tests, with positive RT-LAMP results presenting a characteristic banding pattern on BioAnalyzer readouts, as well as similar melting point curves analysis using Realtime PCR.

**Table 4:** Validation Results

|  | **RT-PCR** | **RT-LAMP** |
| --- | --- | --- |
| **Specificity** | 100%<br>(168/168) | 100%<br>(168/168) |
| **Sensitivity** | 85.5%<br>(65/76) | 78.9%<br>(60/76) |
| **PPV** | 100%<br>(65/65) | 100%<br>(60/60) |
| **NPV** | 93.9%<br>(168/179) | 91.3%<br>(168/184) |

Differences between RT-PCR and RT-LAMP results are not statistically significant (p = 0.44) and the proportion of agreement was 0.89. Kappa value is 0.71.

## V. DISCUSSION

Efforts to control SARS-CoV-2 depend on sensitive (>70%), highly specific (>98%), fast turnaround time diagnostic tests. (Larremore et al. 2020) In this study we describe a scalable method that is accurate, time and cost effective, and displays a sensitivity and specificity similar to the gold standard RT-qPCR. As a result, it has enormous potential in helping to identify infected individuals to control disease spread.

Several molecular detection kits have been developed for faster and more accurate detection of SARS-CoV-2 virus. As expected, sensitivity has been shown to vary according to the sample analyzed. RT-LAMP assays with viral RNA extracted from swab samples demonstrated high sensitivity ranging from 94 to 100%. (https://www.fda.gov/media/138249/download) However, it is not clear if the sensitivity of RT-LAMP from nasopharyngeal swabs can be comparable to RT-LAMP using saliva specimens. Our direct RT-LAMP test from saliva samples has a lower sensitivity as compared to RT-PCR from nasopharyngeal swabs (78.9% *vs*. 85.5%), an observation also made by another saliva-direct RT-LAMP study. (Bhadra et al. 2020)

Noteworthy, some SARS-CoV-2 positive samples were only identified (and confirmed) by RT-LAMP. Considering we were testing different biological samples, this result may be attributed to the presence of the virus solely in saliva of infected patients. The comparison of different techniques, RT-LAMP and RT-PCR, in different types of biological samples is a limitation of this work. This comparison may render incongruent results due to the presence of virus in only one of these biological samples, and not represent the real efficiency of the methods.

The protocol described and validated here focuses on scalability and short turnaround times. Variations on this protocol, such as the addition of standard RNA extraction and concentration can be used to improve sensitivity in detriment of speed and cost. Alternatively, increase in the fluorescence detection cutoff, with removal of the dissociation curve, can be used to speed and decrease costs in detriment of sensitivity and specificity. Each variation can serve distinct purposes (screening or diagnosis), fit different geographic location socio-economic constraints, and moment of the pandemic.

In conclusion, we contribute efforts to contain the pandemic by describing and validating a scalable, fast, and low cost SARS-CoV-2 test that can be easily collected. The ease of implementation should allow this to be implemented in both developed and developing countries.

## Data Availability

Data available upon request.

